# High plasma D-dimer levels correlate with ictal infarction and poor outcomes in spontaneous subarachnoid hemorrhage

**DOI:** 10.1101/2024.05.26.24307957

**Authors:** Hitoshi Kobata, Akira Sugie, Adam Tucker, Gemmalynn Sarapuddin, Hitomi Kimura, Hitoshi Takeshita, Munenori Morihara, Makiko Kawakami

**Affiliations:** Osaka Mishima Emergency Critical Care Center, Takatsuki, Japan; Department of Neurosurgery/Emergency and Critical Care Medicine, Osaka Medical and Pharmaceutical University; Emergency Medical Center, Ijinkai Takeda General Hospital; Department of Neurosurgery, Japanese Red Cross Kitami Hospital; Neurology Department, Institute of Neurosciences, The Medical City; Osaka Medical and Pharmaceutical University Mishima-Minami Hospital; Kyoto Tachibana University; Osaka Mishima Emergency Medical Center; Department of Anesthesiology, Osaka Saiseikai Suita Hospital

**Keywords:** D-dimer, early brain injury, ictal infarction, magnetic resonance imaging, outcome, subarachnoid hemorrhage

## Abstract

**BACKGROUND:** Early brain injury (EBI) is the leading cause of poor outcomes in patients with spontaneous subarachnoid hemorrhage (sSAH). Plasma D-dimer levels and acute cerebral ischemia have been highlighted as relevant findings in EBI; however, their correlation has not been substantially investigated.

**METHODS:** This retrospective, single-center cohort study was conducted at a tertiary emergency medical center from January 2004 to June 2022. Consecutive patients with sSAH who presented within 12 h of SAH ictus and underwent magnetic resonance imaging within 3 days were included. We assessed the correlation of plasma D-dimer levels with acute ischemic lesions detected on the diffusion-weighted images (DWI) and the clinical characteristics of patients.

**RESULTS:** Among 402 eligible patients (mean age, 63.5 years; 62.7% women; median time from onset to arrival, 45.5 min), 140 (34.8%) had acute ischemic lesions. Higher plasma D-dimer levels linearly correlated with worse neurological grades, more severe SAH on initial CT, acute ischemic lesions, and poor outcomes, except for patients with neurogenic stunned myocardium. In the multivariate analysis, acute ischemic lesions were significantly associated with worse neurological grades, higher plasma D-dimer levels, bilateral loss of light reaction, and advanced age. The receiver operating characteristic curve analysis showed D-dimer levels as excellent predictors for acute ischemic lesions (area under the curve [AUC], 0.897; cut-off value, 5.7 µg/mL; p <0.0001) and unfavorable outcomes (AUC, 0.786; cut-off value, 4.0 µg/mL; p <0.0001).

**CONCLUSIONS:** High plasma D-dimer levels correlated with the appearance of acute ischemic lesions on DWI and were dose-dependently associated with worse neurological grades, more severe hemorrhage, and worse outcomes. Plasma D-dimer levels provide a valuable perspective on various issues in the acute phase of sSAH.

## Introduction

Early brain injury (EBI) has emerged as the leading cause of poor outcomes in patients with spontaneous subarachnoid hemorrhage (sSAH).^1,2^ EBI refers to a cascade of brain injuries caused by multiple mechanisms that develop within 72 h of SAH onset. The clinical research interest in sSAH has shifted from delayed cerebral ischemia (DCI) to EBI.^1,2^ The initial events of EBI include global cerebral ischemia and mechanical damage driven by extravasated blood at ictus. Intracranial circulatory arrest may occur in severe cases,^3^ producing profound global cerebral ischemia and subsequent reperfusion injury.

Widespread platelet aggregation develops in cerebral microvessels minutes after ictus and, together with vasoconstriction, induces microvascular perfusion deficits.^4.5^ The transient cessation of cerebral blood flow clinically manifests as loss of consciousness (LOC)^6^ and causes acute ischemic lesions on computed tomography (CT)^7,8^ or diffusion-weighted imaging (DWI) in magnetic resonance imaging (MRI).^9–13^ This “ictal infarction” is related to poor neurological status and unfavorable outcomes.

D-dimer is a soluble degradation product of cross-linked fibrin generated during fibrinolysis and may increase with various illnesses and physiological conditions.^14^ In sSAH, elevated plasma D-dimer levels may reflect the extent of microcirculatory failure,^15^ occur more frequently in poor-grade patients, and predict unfavorable outcomes.^16–20^

Both D-dimer levels and acute ischemic lesions are linked to EBI and unfavorable outcomes; however, this link has not been substantially explored. We aimed to elucidate the association between plasma D-dimer levels, ictal infarction, and outcomes. Furthermore, we discussed the clinical issues associated with hyperacute sSAH from the viewpoint of plasma D-dimer levels.

## Methods

### Ethics statement

This study was approved by the ethics committee of the Osaka Mishima Emergency Critical Care Center. The need for informed consent was waived owing to the retrospective review of unidentified data. All procedures in this study were conducted in accordance with the ethical standards defined in the 1964 Declaration of Helsinki and its later amendments or comparable ethical standards.

### Study design and population

This retrospective, single-center cohort study was conducted at a tertiary emergency medical center. We analyzed 681 consecutive patients with sSAH prospectively registered in the SAH database of our facility between January 2004 and June 2022. SAH was diagnosed based on initial head CT findings. CT angiography (CTA), digital subtraction angiography (DSA), or both were used to identify the source of bleeding in an emergency setting. All patients with sSAH were eligible, including ruptured cerebral aneurysms, arterial dissecting lesions, or unknown causes but not traumatic origin. Patients successfully resuscitated from cardiac arrest (CA) were also included. The exclusion criteria were: 1) arrival >12 h after SAH onset or unidentified onset time (57 patients); 2) no MRI performed (164 patients); 3) MRI performed on day 4 or later (16 patients); and 4) acute ischemic lesions appearing in the perfusion territories potentially attributable to surgical or endovascular procedures (42 patients). Consequently, 402 patients were included (Figure S1).

### Treatment protocol

Comatose patients with unstable cardiopulmonary function were immediately resuscitated in the emergency room. Patients were intubated with muscle relaxants after thorough analgesia and sedation, with a target systolic blood pressure (BP) of 120 mmHg. Nicardipine was added intravenously if necessary. The attending cardiologist performed echocardiography during the initial resuscitation. The patients underwent blood sampling, followed by chest radiography and head CT. When SAH was detected, CTA was subsequently performed, except in patients who remained in CA. Neurological grades were assessed using the Glasgow Coma Scale (GCS) and categorized according to the World Federation of Neurological Surgeons (WFNS)^21^ and modified WFNS^22^ grading systems (Table 1). The CT findings were classified using the modified Fisher CT scale.^23^

**Table 1.** The WFNS and modified WFNS grades for SAH.

| WFNS grade |  |  | modified WFNS grade |  |
| --- | --- | --- | --- | --- |
| Grade | GCS score | Motor deficit | Grade | GCS score |
| I | 15 | absent | I | 15 |
| II | 14-13 | absent | II | 14 |
| III | 14-13 | present | III | 13 |
| IV | 12-7 | present or absent | IV | 12-7 |
| V | 6-3 | present or absent | V | 6-3 |

GCS indicates Glasgow Coma Scale; WFNS, World Federation of Neurosurgeons.

Sources of bleeding were treated immediately after diagnosis as much as possible, primarily through surgical clipping. However, securing the bleeding source was withheld for grade V patients with persistently fixed and dilated pupils after resuscitation. When radical treatment was indicated for grade V patients, therapeutic hypothermia at 34 ℃ was initiated for patients aged ≤75, as described previously.^24^ A cisternal or ventricular drain was placed to control intracranial pressure (ICP) and facilitate the drainage of bloody cerebrospinal fluid. The bone flap was left removed for grade V patients. Patients with arterial dissecting lesions underwent surgical or endovascular trapping, with revascularization if indicated. Patients were managed based on the SAH treatment guidelines of the Neurocritical Care Society and Japan Stroke Society.

### D-dimer measurement

Plasma D-dimer levels were measured by Latex agglutination assay using Nanopia^®^ D-dimer (Sekisui Medical Co., Ltd., measurement range, 0.5–60 μg/mL; normal range, <1.0 μg/mL).

### MRI assessment

Patients underwent an initial MRI study using the 1.5 Tesla EXCELART XG system (Toshiba Medical Systems Co., Tochigi, Japan), principally on postoperative day 1 after securing the bleeding source. Patients who did not receive aggressive treatment for sSAH underwent MRI whenever their hemodynamics stabilized.

Three authors (A. S., H. Kimura, and H. Kobata independently assessed the MRI findings by concealing the blood analysis data. Acute ischemic lesions of >10 mm^2^, depicted by DWI, were considered positive; spotty lesions of ≤10 mm^2^ were considered negative.^10^

### Outcome assessment

The outcome was assessed using the modified Rankin Scale (mRS) at 6 months and dichotomized into favorable (scores 0–2) and unfavorable (sores 3–6). Patients who were discharged alive were referred from rehabilitation hospital reports. As the clinical diagnosis of DCI may be unfeasible in comatose or sedated patients, we assessed the appearance of delayed cerebral infarction on CT or MRI.

### Statistical analysis

Statistical analyses were performed using JMP software (version 17.0.0; SAS Institute Inc., Cary, NC, USA), and a two-sided p <0.05 was considered significant. The baseline characteristics were summarized as numbers (%) for categorical variables and compared using the Chi-square test or Fisher’s exact test, as appropriate. Continuous variables were compared using Student’s *t*-test or Wilcoxon/Kruskal–Wallis test based on the normality of the distribution and are presented as means ± standard deviations or medians (interquartile ranges). Stepwise multivariate logistic regression models were built to analyze the association between D-dimer levels and early ischemic lesions and outcomes. Clinical variables with p <0.25 in the univariate analysis were selected, excluding possible confounding factors. The entry and exit criteria for the model were 0.25 and 0.1, respectively. Receiver operating characteristic (ROC) curve analysis was performed to estimate the sensitivity and specificity of D-dimer levels for acute ischemic lesions and outcomes. The area under the curve (AUC) was calculated to measure the predictive ability.

## Results

### Baseline characteristics

The baseline characteristics of the eligible patients are summarized in Table 2. The mean age was 63.5 ± 12.7 years, with a predominance of women (62.7%). The median time from onset to arrival was 45.5 min. Nearly half of the patients (44.8%) had GCS scores of ≤6, and 13 (3.2%) were resuscitated from CA. The light reaction was lost bilaterally in 29.1% and unilaterally in 0.7% of patients. The pupil diameter was ≥5 mm bilaterally in 7.4% and unilaterally in 4.0% of patients. Neurogenic stunned myocardium and pulmonary edema were present in 13.2% and 11.7% of patients, respectively. Most patients had severe SAH, with grade 3 (57.1%) or 4 (20.9%) on the modified Fisher CT scale. After excluding 19 patients with undetected bleeding sources, SAH derived from lesions in the posterior circulation in 71 (18.5%) patients. Of 42 (11.0%) arterial dissecting lesions, 36 were in the vertebrobasilar arteries, accounting for 50.7% of all lesions in the posterior circulation. Ten of the 19 patients with unknown bleeding sources showed perimesencephalic distribution of SAH. Most patients underwent MRI the day after treating the bleeding source, i.e., day 1 of SAH. The DWI revealed acute ischemic lesions in 140 (34.8%) patients.

**Table 2.**
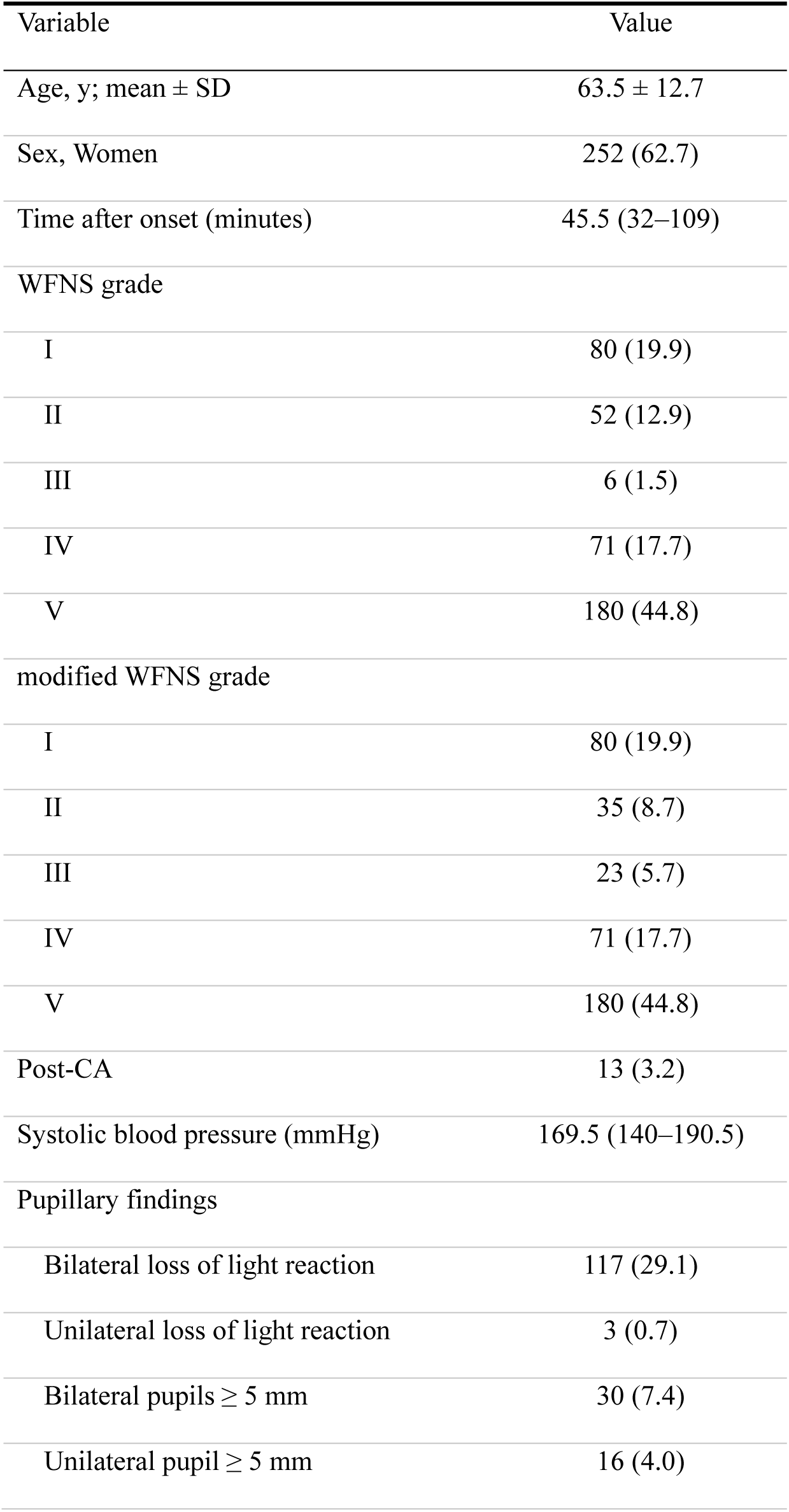
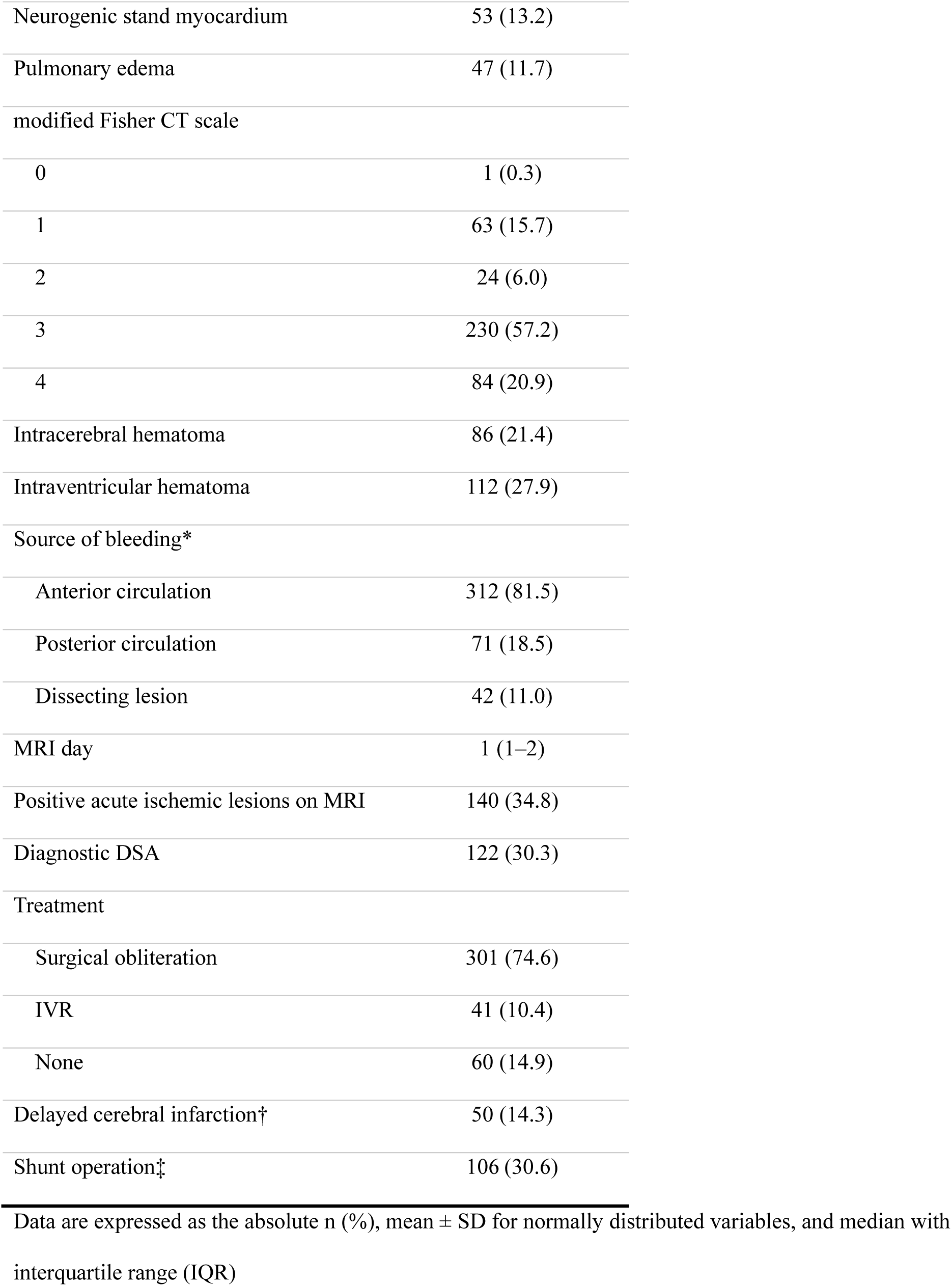
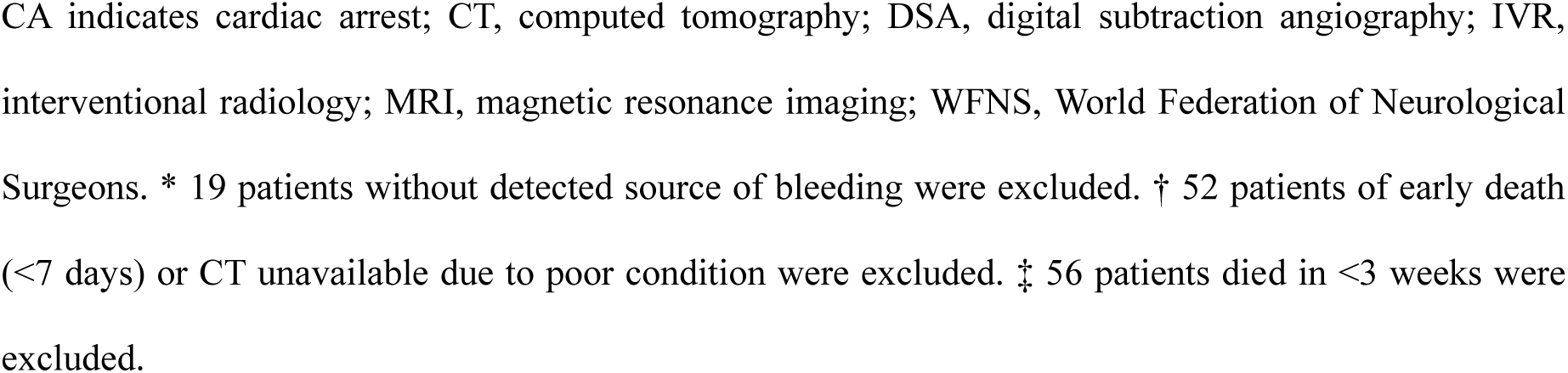
Baseline Characteristics of Patients.

| Variable | Value |
| --- | --- |
| Age, y; mean ± SD | 63.5 ± 12.7 |
| Sex, Women | 252 (62.7) |
| Time after onset (minutes) | 45.5 (32–109) |
| WFNS grade |  |
| I | 80 (19.9) |
| II | 52 (12.9) |
| III | 6 (1.5) |
| IV | 71 (17.7) |
| V | 180 (44.8) |
| modified WFNS grade |  |
| I | 80 (19.9) |
| II | 35 (8.7) |
| III | 23 (5.7) |
| IV | 71 (17.7) |
| V | 180 (44.8) |
| Post-CA | 13 (3.2) |
| Systolic blood pressure (mmHg) | 169.5 (140–190.5) |
| Pupillary findings |  |
| Bilateral loss of light reaction | 117 (29.1) |
| Unilateral loss of light reaction | 3 (0.7) |
| Bilateral pupils ≥ 5 mm | 30 (7.4) |
| Unilateral pupil ≥ 5 mm | 16 (4.0) |
| Neurogenic stand myocardium | 53 (13.2) |
| Pulmonary edema | 47 (11.7) |
| modified Fisher CT scale |  |
| 0 | 1 (0.3) |
| 1 | 63 (15.7) |
| 2 | 24 (6.0) |
| 3 | 230 (57.2) |
| 4 | 84 (20.9) |
| Intracerebral hematoma | 86 (21.4) |
| Intraventricular hematoma | 112 (27.9) |
| Source of bleeding* |  |
| Anterior circulation | 312 (81.5) |
| Posterior circulation | 71 (18.5) |
| Dissecting lesion | 42 (11.0) |
| MRI day | 1 (1–2) |
| Positive acute ischemic lesions on MRI | 140 (34.8) |
| Diagnostic DSA | 122 (30.3) |
| Treatment |  |
| Surgical obliteration | 301 (74.6) |
| IVR | 41 (10.4) |
| None | 60 (14.9) |
| Delayed cerebral infarction† | 50 (14.3) |
| Shunt operation‡ | 106 (30.6) |

Data are expressed as the absolute n (%), mean ± SD for normally distributed variables, and median with interquartile range (IQR)
CA indicates cardiac arrest; CT, computed tomography; DSA, digital subtraction angiography; IVR, interventional radiology; MRI, magnetic resonance imaging; WFNS, World Federation of Neurological Surgeons. \* 19 patients without detected source of bleeding were excluded. † 52 patients of early death (<7 days) or CT unavailable due to poor condition were excluded. ‡ 56 patients died in <3 weeks were excluded.

Diagnostic DSA was performed in 122 (30.3%) patients. The bleeding sources were secured in 342 patients, with surgical clipping in 288 (71.6%), trapping in 13 (3.2%), and IVR in 41 (10.2%) patients, excluding 60 (14.9%) with devastating conditions or unknown causes of SAH. Delayed cerebral infarction was identified in 50 (14.3%) patients, excluding 52 early deaths and those unavailable for examination. Shunt surgery was performed on 105 (30.4%), excluding 56 patients who died <3 weeks.

### Univariate analyses for acute ischemic lesions and outcomes

Univariate analyses for the associated variables with acute ischemic lesions on DWI and outcomes are summarized in Table S1. The appearance of acute ischemic lesions was significantly associated with the following: older age; more severe neurological grade based on WFNS and modified WFNS grading systems; bilateral loss of light reaction and pupillary dilatation; higher blood glucose levels, glucose/potassium ratio, and D-dimer levels; prolonged prothrombin time and activated partial thromboplastin time; decreased platelet counts; increased white blood cell (WBC) counts; more severe modified Fisher CT grade; the association of intracerebral hematoma (ICH) or intraventricular hematoma (IVH); lower incidence of DSA performance; a bleeding source in the posterior circulation; neurogenic stunned myocardium; pulmonary edema; delayed cerebral infarction; and subsequent shunt operation for chronic hydrocephalus. No significant differences were identified in sex, time after onset to arrival, serum potassium and fibrinogen levels, C-reactive protein, arterial dissecting lesions, and treatment procedures.

The relevant factors for outcomes were similar to those for acute ischemic lesions; however, lower systolic BP and dissecting lesions were included for favorable outcomes, and platelet counts and neurogenic stunned myocardium were excluded.

Sixteen patients presenting with unilateral pupillary dilation had a higher incidence of ipsilateral ICH (43.8% vs. 20.8%, p=0.026) and aneurysms in the internal carotid-posterior communicating artery or middle cerebral artery (75% vs. 25%, p=0.0038), compared with patients without this abnormality.

### Univariate analysis for D-dimer levels and clinical characteristics

Table 3 presents the plasma D-dimer levels and clinical characteristics. Higher D-dimer levels were associated with worse WFNS and modified WFNS grades, abnormal pupillary findings, worse modified Fisher CT scale grades, association of ICH or IVH, neurogenic stunned myocardium, pulmonary edema, acute cerebral ischemic lesions, shunt operation, delayed cerebral infarction, and worse outcomes. D-dimer levels showed no significant differences by the bleeding sources, either anterior or posterior circulation, and saccular aneurysm or dissecting lesion.

**Table 3.**
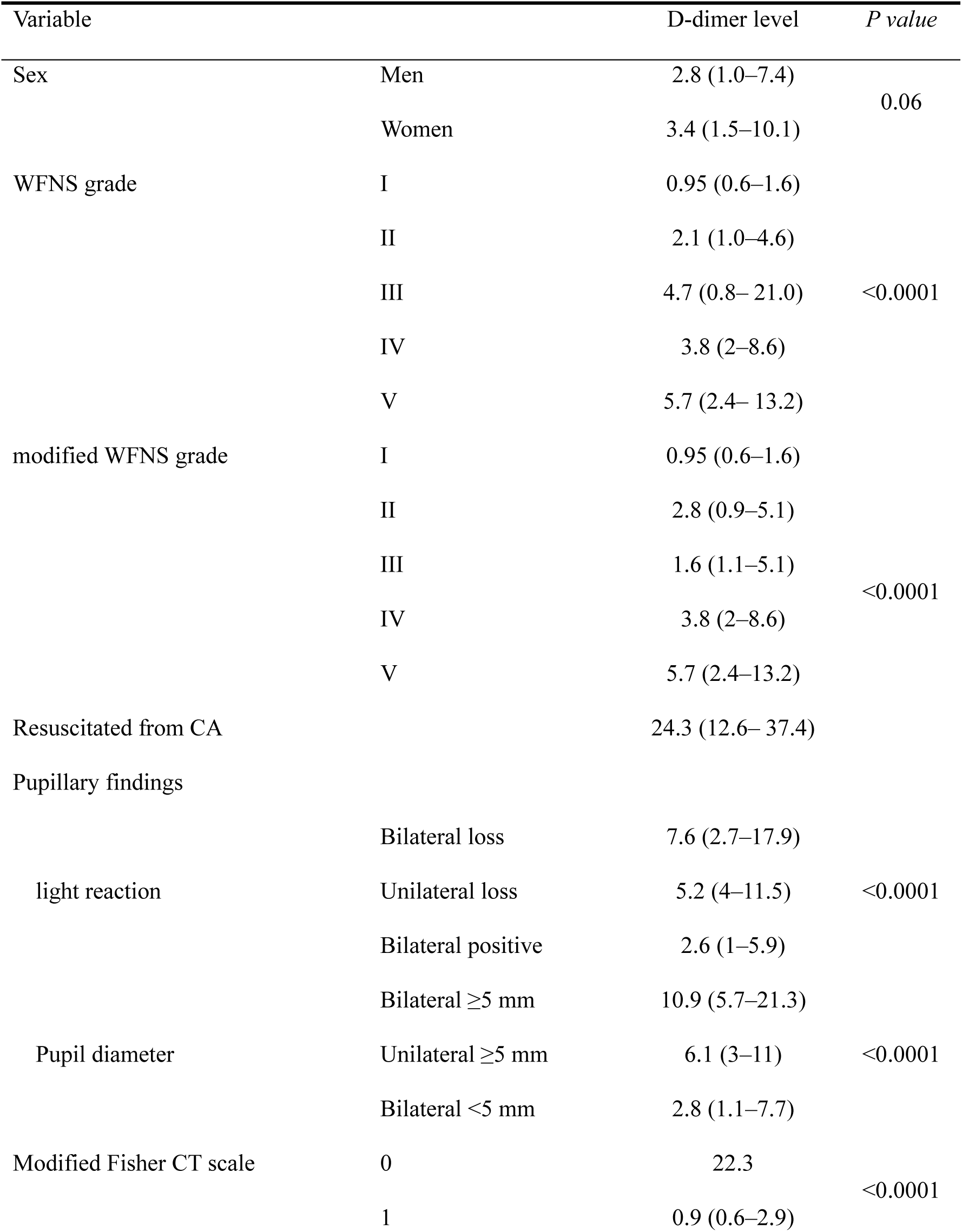
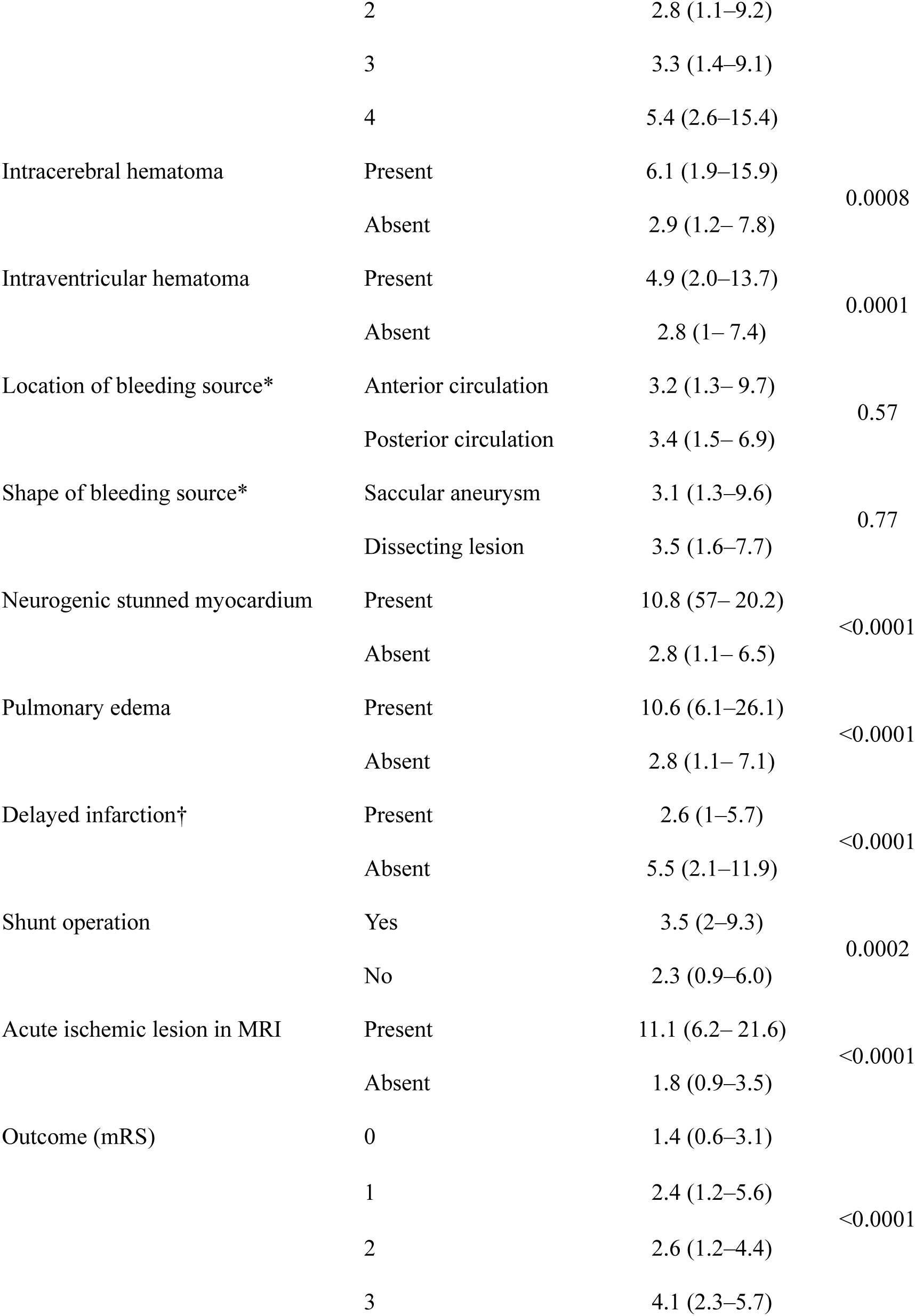
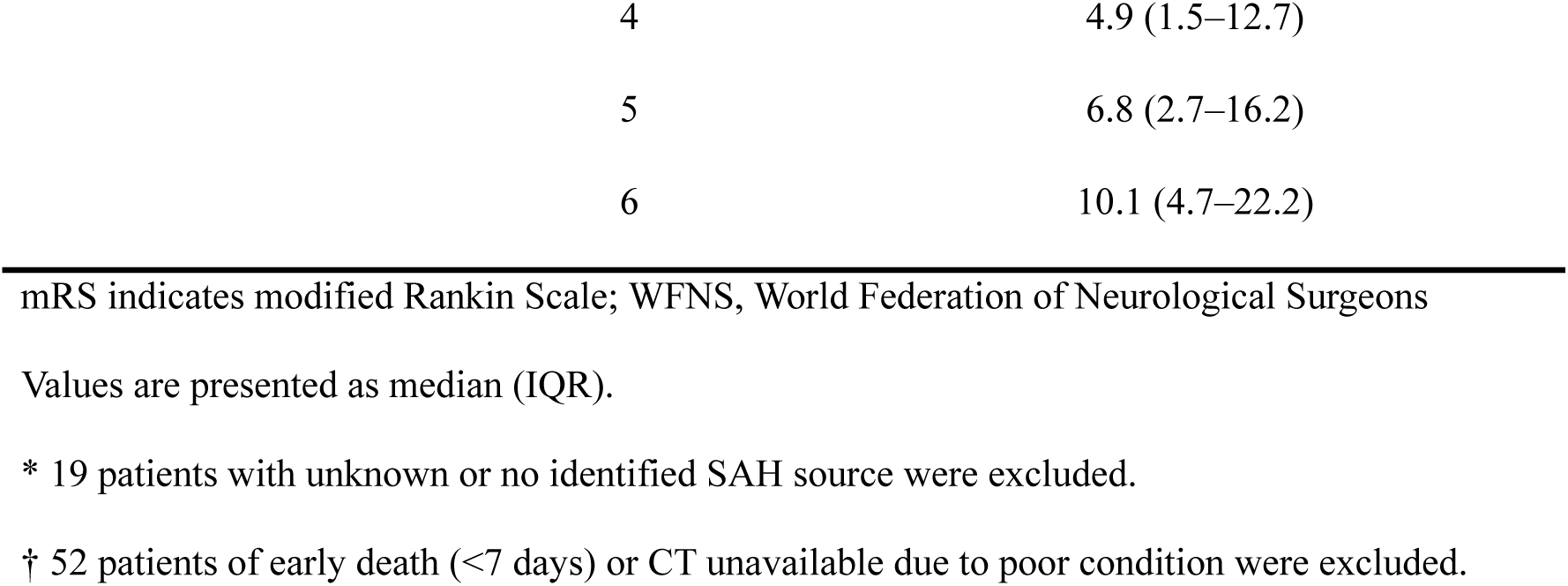
Univariate Analysis of D-dimer Levels and Clinical Characteristics.

Trends for higher D-dimer levels were observed as the neurological grade worsened; however, no significant differences were observed between grades II and III, and III and IV in the WFNS grade, and between grades II and III in the modified WFNS grade (Fig. 1A and B) (p <0.0001).

**Figure. 1.**
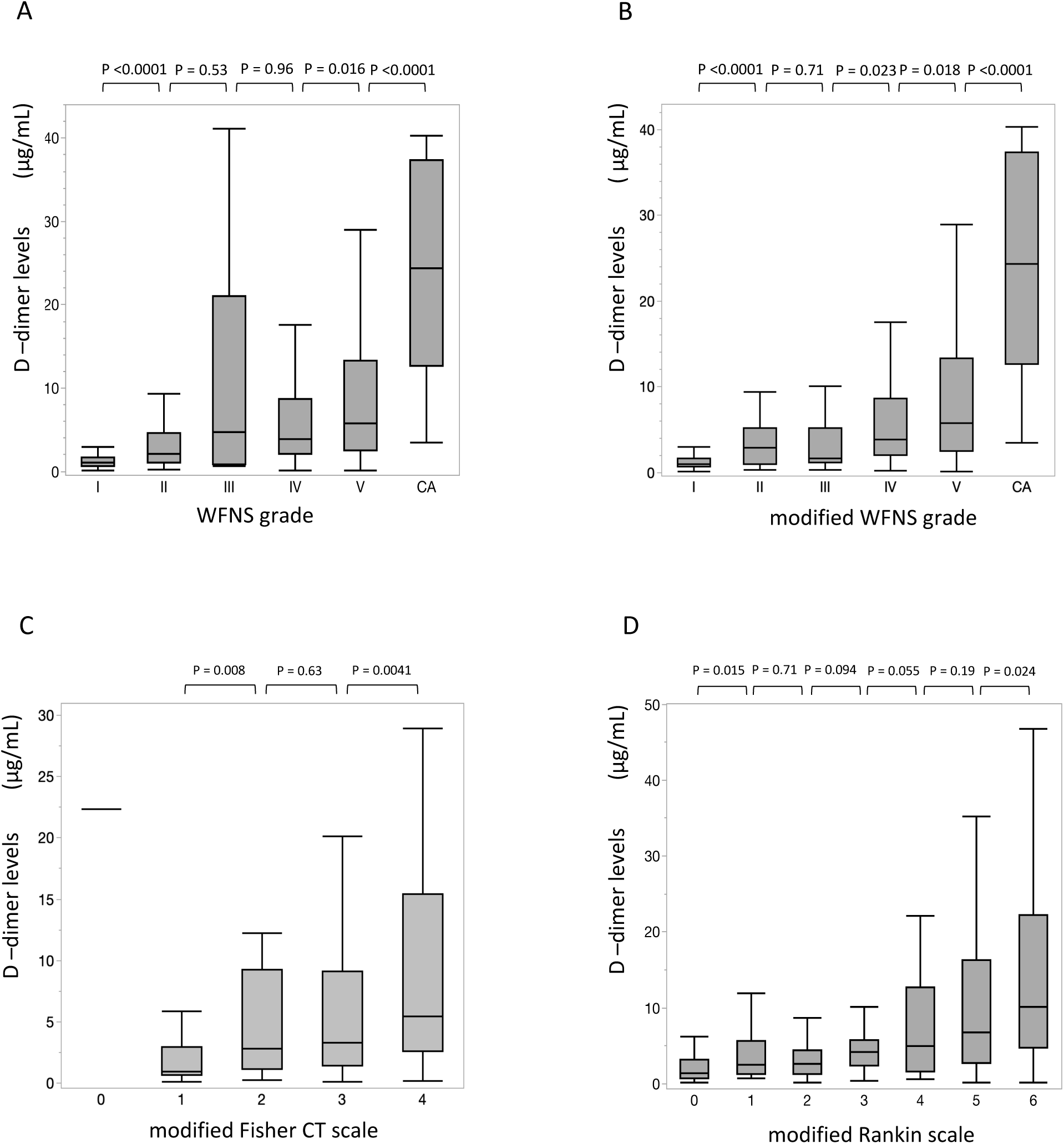
Association of serum D-dimer levels with neurological severity, CT findings at presentation, and outcomes. Higher plasma D-dimer levels are associated with worse WFNS (A) and modified WFNS (B) grades (including post-cardiac arrest), modified Fisher CT scale (C), and modified Rankin scale (D) (p <0.0001). P values between each group are shown. WFNS indicates the World Federation of Neurological Surgeons.

Higher D-dimer levels were associated with more severe SAH assessed by the modified Fisher CT scale (p <0.0001) (Fig. 1C). A grade 0 patient with high D-dimer level had no SAH but acute subdural hematoma causing cerebral herniation. D-dimer levels were significantly lower in patients with perimesencephalic non-aneurysmal SAH than those in patients whose bleeding sources were identified (Figure S2). A linear trend toward worse outcomes was identified with higher D-dimer levels (Fig. 1D) (p <0.0001). The difference was significant between 0 and 1, and between 5 and 6 for each mRS score.

### Association of neurological grades and acute ischemic lesions

A trend toward an increased occurrence of acute ischemic lesions was observed as the WFNS and modified WFNS grades worsened (Figure S3A and S3B) (p <0.0001). Exceptionally, several good-grade patients with dissecting lesions simultaneously developed SAH and acute ischemia in the vascular territories distal to the involved lesion.

### Multivariate analysis of associated variables for acute ischemic lesions and outcomes

In the multivariate analysis, worse modified WFNS grades, higher plasma D-dimer levels, bilateral loss of light reaction, and advanced age were significant predictors of acute ischemic lesions. In addition, modified Fisher CT grades and WBC counts were significant outcome predictors (Table 4).

**Table 4.**
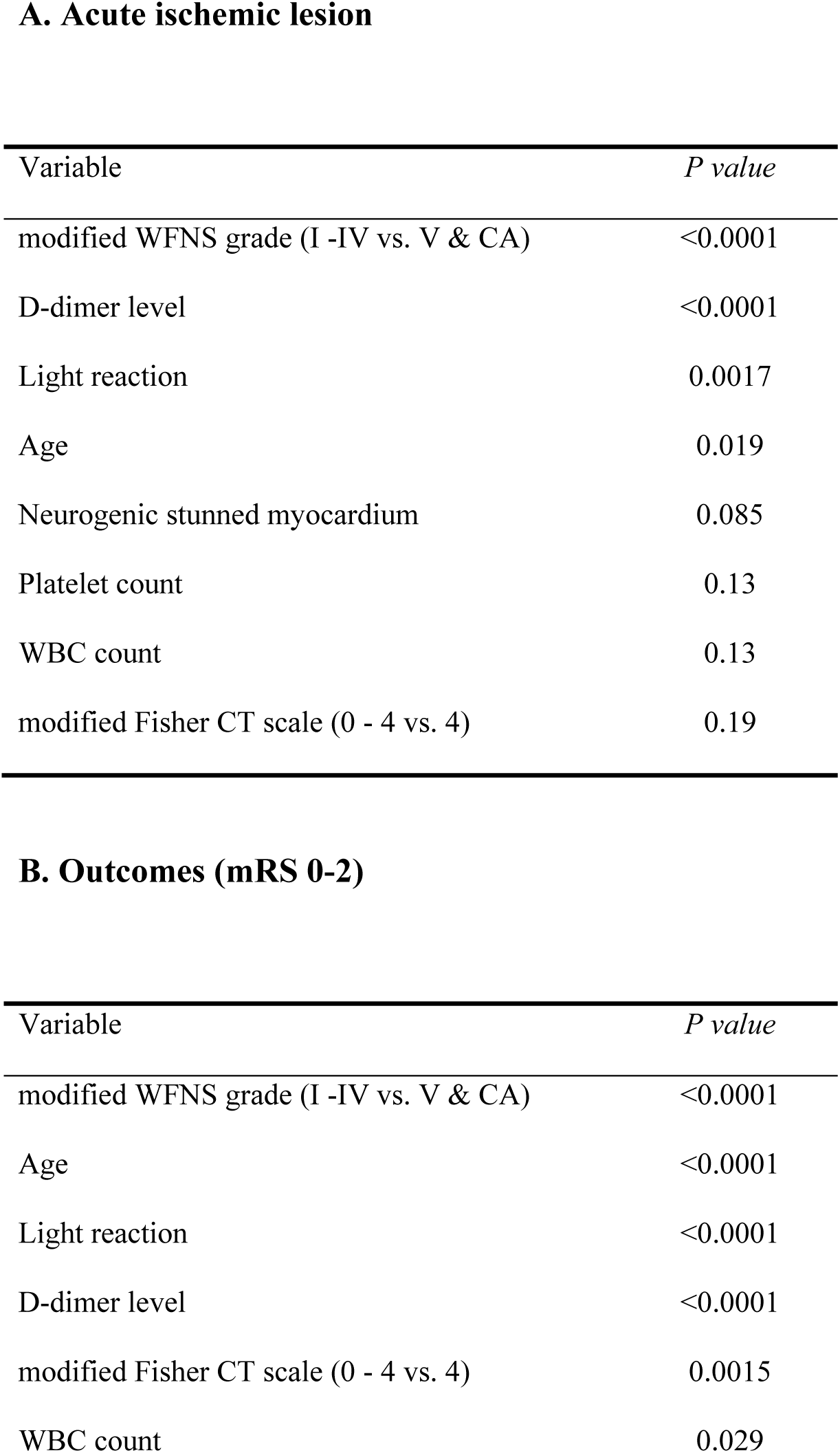
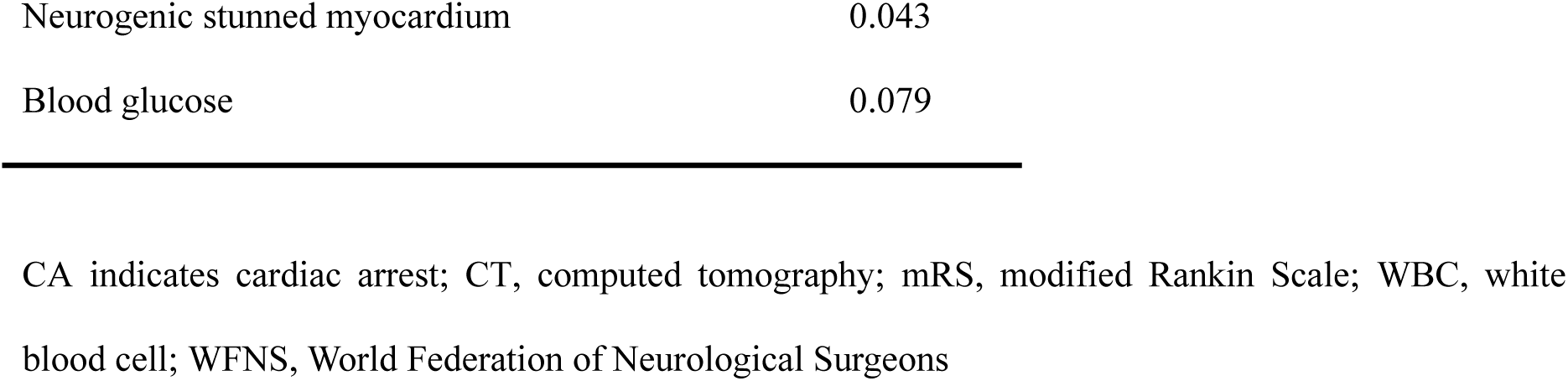
Multivariate Analysis for Factors Associated with Acute Ischemic Lesions (A) and Outcomes (B)

**A. Acute ischemic lesion**
| Variable | *P value* |
| --- | --- |
| modified WFNS grade (I -IV vs. V & CA) | <0.0001 |
| D-dimer level | <0.0001 |
| Light reaction | 0.0017 |
| Age | 0.019 |
| Neurogenic stunned myocardium | 0.085 |
| Platelet count | 0.13 |
| WBC count | 0.13 |
| modified Fisher CT scale (0 - 4 vs. 4) | 0.19 |

**B. Outcomes (mRS 0-2)**
| Variable | *P value* |
| --- | --- |
| modified WFNS grade (I -IV vs. V & CA) | <0.0001 |
| Age | <0.0001 |
| Light reaction | <0.0001 |
| D-dimer level | <0.0001 |
| modified Fisher CT scale (0 - 4 vs. 4) | 0.0015 |
| WBC count | 0.029 |
| Neurogenic stunned myocardium | 0.043 |
| Blood glucose | 0.079 |

CA indicates cardiac arrest; CT, computed tomography; mRS, modified Rankin Scale; WBC, white blood cell; WFNS, World Federation of Neurological Surgeons

### ROC analysis of D-dimer levels for acute ischemic lesions and outcomes

The ROC curve analysis showed D-dimer levels as excellent predictors for acute ischemic lesions (AUC, 0.897; cut-off value, 5.7 µg/mL; p < 0.0001) (Fig. 2A) and unfavorable outcomes (AUC, 0.786; cut-off value, 4.0 µg/mL; p < 0.0001) (Fig. 2B).

**Figure. 2.**
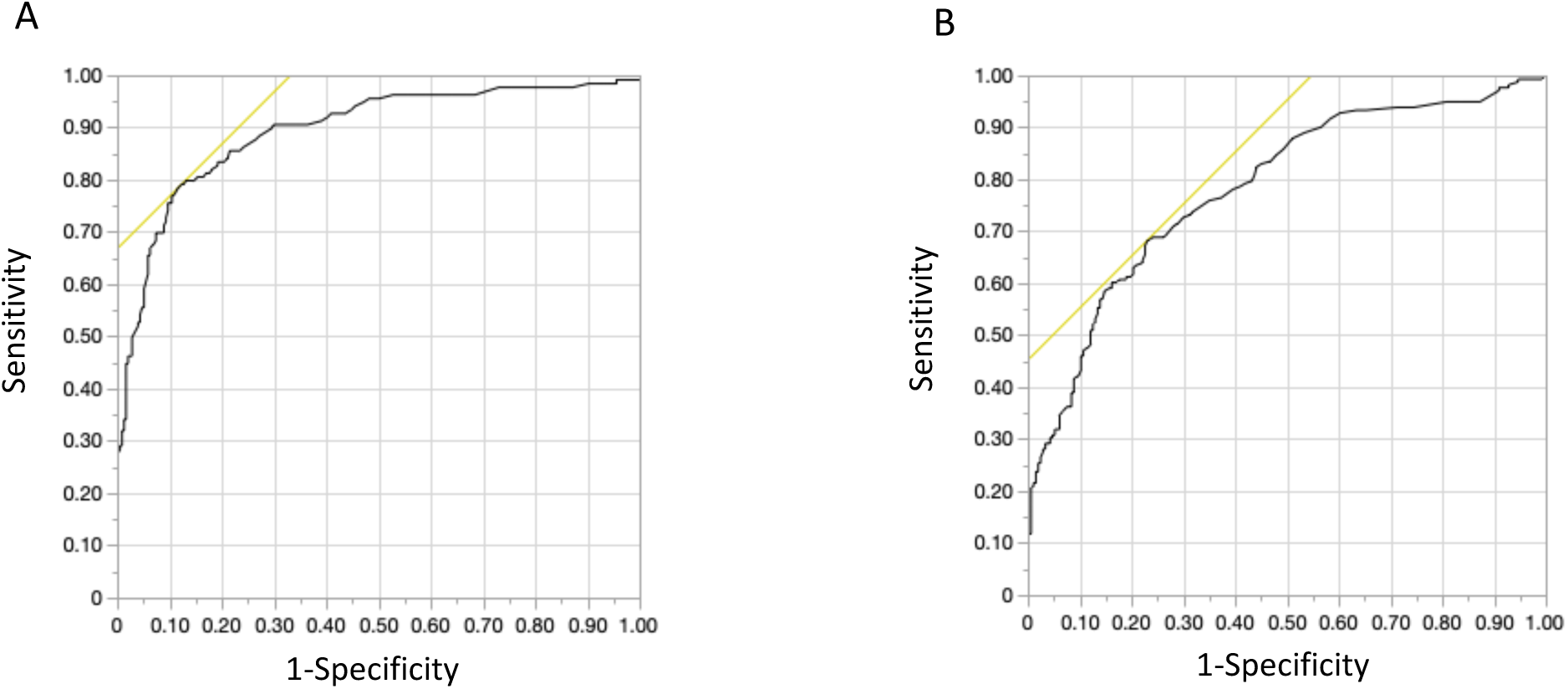
Receiver operating characteristic curve (ROC) comparison of D-dimer levels with acute ischemic lesions and outcomes. The ROC curve analyses show that higher D-dimer levels as excellent predictors for acute ischemic lesions (A) (AUC=0.897: The optimal cut-off value = 5.7 µg/mL) and unfavorable outcomes assessed by the modified Rankin scale (B) (AUC=0.786: The optimal cut-off value = 4.0 µg/mL).

### Association of acute cerebral ischemic lesions and outcomes

The appearance of acute ischemic lesions was associated with unfavorable outcomes (Figure S4A). When dichotomized into favorable and unfavorable outcome groups, acute ischemic lesions were significantly associated with unfavorable outcomes (86.4% vs. 13.6%; odds ratio, 20.1; 95% confidence interval, 11.5–35.2) (Figure S4B).

### Subgroup with favorable outcomes without acute ischemic lesions, despite high D-dimer levels

Of 24 patients with D-dimer levels above the 90^th^ percentile for early ischemic lesions (>6.07 µg/mL), 11 had neurogenic stunned myocardium. Likewise, of 21 patients with D-dimer levels above the 90^th^ percentile for poor outcomes (>9.33 µg/mL), 14 had neurogenic stunned myocardium. One patient with multiple subcutaneous hemorrhages after repeated fallover after SAH ictus had a favorable outcome although D-dimer exceeded 90^th^ percentile.

## Discussion

High plasma D-dimer levels on admission were associated with the appearance of acute ischemic lesions. Higher D-dimer levels dose-dependently correlated with worse neurological grades, more severe hemorrhage on initial CT, and unfavorable outcomes in patients with sSAH. These associations suggest that D-dimer is an essential indicator in the pathogenesis of EBI. However, in cases of neurogenic stunned myocardium, high D-dimer levels did not always indicate severe EBI.

Intracranial circulatory arrest at SAH onset promotes platelet aggregation in the parenchymal microvasculature minutes after ictus. Platelet counts decline rapidly from minutes to hours, and aggregated platelets may contribute to microvascular perfusion deficits.^4^ In experimental SAH, T2* MRI detects cerebral vessel thrombosis within 4 h^25^ and T2 hyperintensity area matched with fibrinogen/fibrin-positive staining.^26^ With the restoration of cerebral blood flow, thrombolysis occurs, promoting D-dimer production. We observed significantly higher D-dimer levels and lower platelet counts in patients with acute ischemic lesions on DWI. These findings highlight D-dimer as an essential indicator of early ischemia-reperfusion injury contributing to EBI.

Previous studies have reported the correlation of high D-dimer levels with poor-grade SAH, ^16–20^ high ICP,^27^ the amount of SAH on initial CT,^17,19,20^ the occurrence of DCI^17,28^ and systemic complications,^19,20^ and unfavorable outcomes.^16–20^ A higher D-dimer quartile was associated with increased long-term mortality, older age, larger aneurysm size, worse Fisher CT group, worse Hunt and Hess grade, and elevated biomarkers.^20^ We demonstrated a dose-dependent association of D-dimer levels with the WFNS and modified WFNS grades, the modified Fisher CT grade, and outcomes. The highest D-dimer levels in post-CA patients may indicate more severe and systemic ischemia-reperfusion injury.

Since the first MRI-based presentation of acute cerebral ischemia,^9^ the characteristics and clinical significance of these lesions have been investigated. The lesions visible on DWI appear to multiply and are often symmetrical in the bilateral cerebral hemispheres, unrelated to the site of the ruptured aneurysm.^9–13^ Lesions are observed predominantly in poor-grade patients and correlated with the amount of SAH, LOC, global cerebral edema,^12^ global hypoperfusion,^29^ delayed extensive ischemic lesions,^30^ and unfavorable outcomes.^9–12^ A meta-analysis demonstrated lesion incidences of 17% in all analyzed cases, 14% in CT-based studies, and 41% in MRI-based studies.^31^

The presumed pathogenesis of acute ischemic lesions shares many of the common features as that of elevated D-dimer levels: microcirculatory disturbances attributable to the “no-reflow phenomenon”,^9^ acute cerebral vasoconstriction, obstruction of small vessels caused by platelet–leukocyte activation,^12^ or decreased cerebral perfusion from neurogenic cardiac dysfunction or pulmonary edema.^11^

Although a small study mentioned an association between D-dimer levels and early ischemic lesions,^13^ this association has not been substantially discussed. We demonstrated a strong correlation between D-dimer levels and acute cerebral ischemic lesions, with a cut-off value of 5.7 µg/mL. The optimal cut-off value for D-dimer for long-term outcomes was reported 2.36 µg/mL.^20^ In our study, the cut-off value of D-dimer for poor outcomes was 4.0 µg/mL.

Plasma D-dimer levels provide valuable insights into clinical issues regarding sSAH. As WFNS grade III is not consistently associated with outcomes, the modified WFNS was proposed, which assigned total GCS scores of 14 and 13 to grades II and III, respectively, regardless of neurological deficits.^22^ This modification yielded significant differences in the outcomes for each grade. Our study showed that focal neurologic symptoms of WFNS grade III were associated with ICH and elevated D-dimer levels. D-dimer levels presented better discrimination between adjacent grades in the modified WFNS than those in the WFNS grading system.

We confirmed that neurogenic stunned myocardium and pulmonary edema are risk factors for ictal infarction, as previously reported.^11^ However, several patients with neurogenic stunned myocardium had favorable outcomes without acute ischemic lesions despite high D-dimer levels. The D-dimer may be of cardiac origin, as high D-dimer levels have been reported in Takotsubo cardiomyopathy.^32^ Favorable outcomes can be expected when satisfactory cerebral circulation is maintained for these patients.

The GCS-Pupil score, which adds pupil reactivity to the GCS, has been reported for the prognostication of patients with traumatic brain injury.^33^ In patients with sSAH, pupillary findings also enhance predictive accuracy.^34,35^ The loss of pupillary reaction in sSAH generally occurs bilaterally due to global cerebral ischemia including the midbrain, where the oculomotor nuclei are located. In contrast, unilateral pupillary dilation is relatively uncommon and could be attributed to direct compression by an aneurysm or uncal herniation rather than global cerebral ischemia.

The optimal BP for maintaining cerebral circulation while preventing rebleeding remains uncertain in hyperacute sSAH. The 2023 Guidelines for SAH do not recommend a specific target for BP reduction.^36,37^ We used short-acting neuroprotective anesthetics to lower the systolic BP to 120 mmHg; subsequently, the patients underwent immediate treatment for bleeding sources. This therapeutic strategy did not appear to increase acute ischemic lesions compared with those in the meta-analysis data.^31^

Hyperfibrinolysis, indicated by high D-dimer levels, may be related to rebleeding after SAH.^20^ Conversely, a significant association between elevated D-dimer levels and increased thromboembolic events was reported during endovascular coiling of ruptured aneurysms.^38^ D-dimer levels were lower in patients with perimesencephalic SAH than those in aneurysmal SAH, consistent with clinical findings in previous studies.^39^ High D-dimer levels with delayed cerebral infarction and shunt surgery may be related to the extent of sSAH, which is associated with D-dimer levels.

Various measures have been reported to assess the severity of sSAH. Neurological grading is the mainstay for this purpose; however, determining severity in sedated or comatose patients is challenging in the hyperacute phase. Stress-induced hyperglycemia,^40^ glucose/potassium ratio,^41^ physiological responses,^42,43^ and imaging findings^44^ serve as references for evaluating severity. We demonstrated that D-dimer is an essential indicator for EBI in sSAH, reflecting ischemia-reperfusion injury in the brain. However, D-dimer levels are not disease-specific and may be associated with extracerebral comorbidities, such as cancer, thromboembolism, and recent trauma.^14^ In contrast, low D-dimer levels indicate mild EBI.

### Limitations

This study has several limitations. First, this was a single-center, retrospective study. A potential bias may have been present. Second, most patients underwent MRI after surgical or endovascular treatment. Procedure-related infarction cannot be completely excluded.

Third, unrecognized rebleeding after blood sampling may have affected the analysis. Finally, pathogenesis other than sSAH causing D-dimer elevation cannot be entirely eliminated.

## Conclusions

Plasma D-dimer levels reflect ischemia-reperfusion injury and are associated with ictal infarction in patients with acute sSAH. High D-dimer levels were associated with worse neurological grades, more severe hemorrhage, and worse outcomes. These findings imply that D-dimer is an essential indicator in the pathogenesis of EBI and a reliable guide to treatment, except for patients with neurogenic stunned myocardium. Furthermore, plasma D-dimer levels provide a valuable perspective on various issues during the acute phase of sSAH.

## Data Availability

N/A

## Non-standard Abbreviations and Acronyms

AUC: area under the curve
BP: blood pressure
CA: cardiac arrest
CT: computed tomography
CTA: computed tomography angiography
DCI: delayed cerebral ischemia
DWI: diffusion-weighted image
EBI: early brain injury
ICH: intracerebral hemorrhage
IVH: intraventricular hemorrhage
IVR: interventional neuroradiology
LOC: loss of consciousness
MRI: magnetic resonance imaging
mRS: modified Rankin Scale
ROC: receiver operating characteristic
sSAH: spontaneous subarachnoid hemorrhage
WFNS: World Federation of Neurological Surgeons

## Acknowledgments

We would like to thank Editage (www.editage.com) for English language editing.

## Source of finding

None.

## Supplemental Material

Table S1 Figures S1-S4

